# Long-term SARS-CoV-2 RNA Shedding and its Temporal Association to IgG Seropositivity

**DOI:** 10.1101/2020.06.02.20120774

**Authors:** Vineet Agarwal, AJ Venkatakrishnan, Arjun Puranik, Christian Kirkup, Agustin Lopez-Marquez, Douglas W. Challener, John C. O’Horo, Matthew J. Binnicker, Walter K. Kremers, William A. Faubion, Andrew D. Badley, Amy W. Williams, Gregory J. Gores, John D. Halamka, William G. Morice, Venky Soundararajan

## Abstract

Analysis of 851 COVID-19 patients with a SARS-CoV-2-positive PCR at follow-up shows 99 patients remained SARS-CoV-2-positive after four weeks from initial diagnosis. Surprisingly, a majority of these long-term viral RNA shedders were not hospitalized (61 of 99), with variable PCR Crossing point values over the month post diagnosis. For the 851-patient cohort, the mean lower bound of viral RNA shedding was 17.3 days (SD: 7.8), and the mean upper bound of viral RNA shedding from 668 patients transitioning to confirmed PCR-negative status was 22.7 days (SD: 11.8). Among 104 patients with an IgG test result, 90 patients were seropositive to date, with mean upper bound of time to seropositivity from initial diagnosis being 37.8 days (95%CI: 34.3-41.3). Juxtaposing IgG/PCR tests revealed that 14 of 90 patients are non-hospitalized and seropositive yet shed viral RNA. This study emphasizes the need for monitoring viral loads and neutralizing antibody titers in long-term shedders.

## Introduction

As COVID-19 continues to rage globally with over 11 million confirmed infected individuals and 500,000 deaths^1^, the world is grappling with the dual challenge of stemming the tide of the current pandemic and planning for reopening the economy in the post-COVID phase. Currently, there are over a million patients that have recovered from COVID-19^1^, and some governments have suggested that antibody-based tests in recovered individuals can be used as the basis for an “immunity passport”^2^ to travel or return-to-work assuming that they are protected against re-infection and likely not infectious. However, there are also emerging reports of viral shedding for many days post-recovery, as evidenced from PCR tests on stool samples^3^ and recurrent SARS-CoV-2-positive PCR tests in “cured” patients^4^.

In addition to routine RT-PCR assays that are the gold standard for COVID-19 clinical diagnosis, recent studies have suggested droplet digital PCR (ddPCR) as a more sensitive assay for quantifying viral load in early infection stages^5,6^. The crossing point (Cp) values from SARS-CoV-2 RT-PCR assays has also been correlated with culture positivity to suggest that Cp values above 33-34 may no longer be associated with replication competent virus^7^.

The general lack of understanding of the period of infectivity, viral shedding, and potential for transmission necessitates longitudinal monitoring of viral clearance in COVID-19 patients. Such an analysis has the potential to help inform the immunological basis for rapid viral control and disease progression.

## Results

Between February and June 2020, 131,646 individuals underwent routine SARS-CoV-2 PCR testing at the Mayo Clinic hospitals in Minnesota, Arizona and Florida, and the associated Mayo Clinic Health System (**Figure 1a**). Of these, 27,309 individuals (21%) were subjected to the PCR test more than once (**Figure 1a**). Of all the individuals tested, 5,699 patients tested SARS-CoV-2-positive (henceforth, COVID_pos_) at least once during the study period (**Figure 1b**). The age distributions in the context of hospitalization, intensive care unit (ICU) admission, and mortality status are shown for COVID_pos_ patients in **Figure 1c-f**. Notably, over 50% of the COVID_pos_ patients in this study are in the age group of 0-40. The pattern of increased hospitalization, ICU admissions and death in the elderly compared to the younger populations is consistent with previous studies of COVID-19 patient demographics^8–10^.

**Figure 1.**
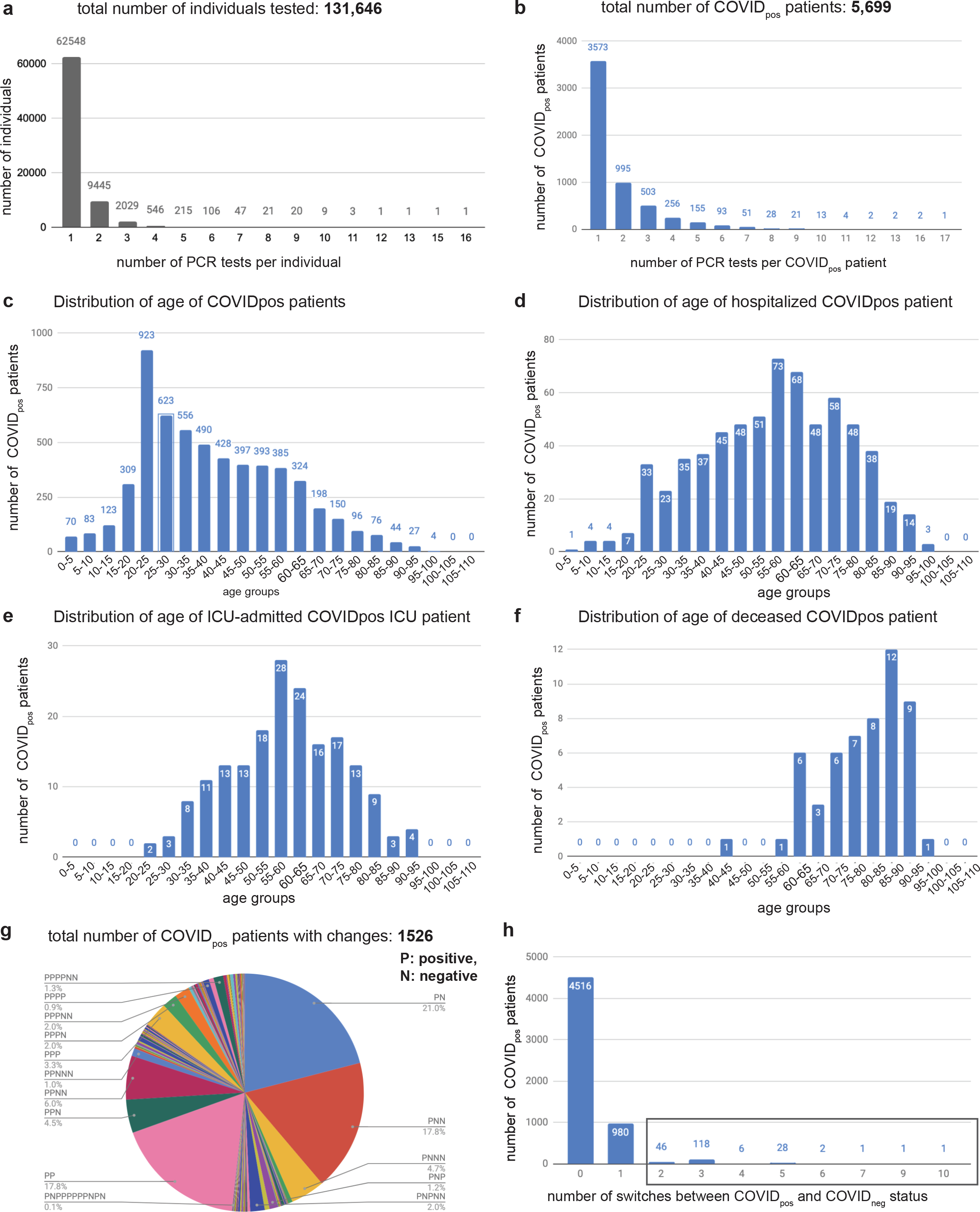
Distributions of **(a)** number of PCR tests per individual, (**b**) number of PCR tests taken by COVID_pos_ patients, (**c**) age of COVID_pos_ patients, (**d**) age of hospitalized COVID_pos_ patients, (**e**) age of ICU-admitted COVIDpos patients, (**f**) age of deceased COVID_pos_ patients, (**g**) the number of patients by sequence of SARS-COV-2 PCR positive and negative results, and (**h**) the number of switches between COVID_pos_ and COVID_neg_ status in longitudinal testing of COVID_pos_ patients; box indicates the count of patients that switched from COVID_pos_ to COVID_neg_ and back to COVID_pos_ status at least once.

Among the 5,699 COVID_pos_ patients, 2,126 patients (37.3%) were subjected to two or more PCR tests, and 851 patients (14.9%) had at least two SARS-CoV-2-positive PCR tests (**Figure 1b**,**g**). We observed 203 patients oscillated from SARS-CoV-2-positive to SARS-CoV-2-negative and back to SARS-CoV-2-positive status one or more times (**Figure 1h**). Given such oscillation potential, we henceforth consider a pair of contiguous SARS-CoV-2 negative PCR tests after the last positive PCR test to be indicative of a ‘confirmed COVID-19 negative’ status.

Despite the caveat of routine RT-PCR assays not providing data on the replication competency of the virus, the availability of these longitudinal PCR test results and the patients’ Electronic Health Records (EHRs), provides an excellent opportunity to quantify the duration of SARS-CoV-2-positive PCR results. Specifically, we aimed to quantify for each patient – (1) a *lower bound of infection duration*, defined as the time in days between the first and last positive SARS-CoV-2 PCR tests, and (2) an *upper bound of infection duration*, defined as the time in days between the first positive PCR test and the second negative PCR test after which there are no further positive PCR tests (**Figure 2a**). The lower bound captures the most intuitive estimate of infection duration, at least from the standpoint of SARS-CoV-2 PCR positivity. Nevertheless, our quantification of the upper bound is motivated by recent reports of high false-negative rates for SARS-CoV-2 PCR tests^11^, our own observation of oscillations in serial PCR testing outcomes (**Figure 1h)**, and the requirement for healthcare workers to observe negative PCR results on two consecutive days to meet the CDC “Return to Work” criteria^8^.

**Figure 2.**
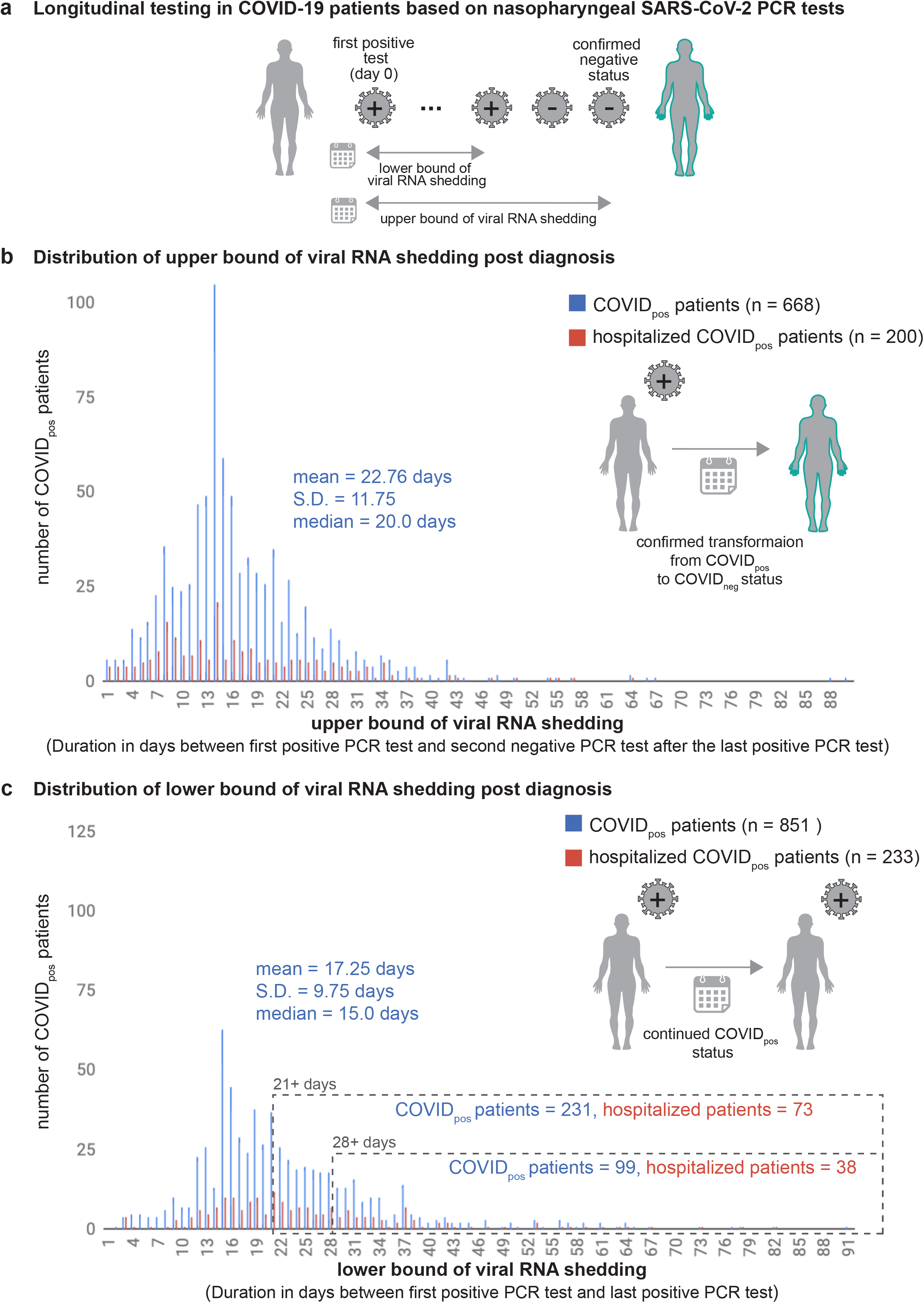
Distribution of the COVID_pos_ patients by **(a)** duration between the day of diagnosis to second contiguous negative test after last positive test. **(b)** duration between the day of diagnosis to the last positive test.

COVID-19 patients whose lower bound of infection duration is greater than four weeks (28 days) are hereafter referred to as patients with ‘long-term shedding of viral RNA’. For the 668 COVID_pos_ patients that switched to a confirmed negative status, i.e. two negative SARS-CoV-2 PCR tests following the last positive SARS-CoV-2 test, the distribution of the upper bound of infection duration was a mean of 22.8 days and a standard deviation of 11.8 days (**Figure 2b**).

Of the 851 COVID_pos_ patients with at least two PCR positive results, interestingly, 231 patients (27%) and 99 patients (11.6%) have viral RNA shedding beyond 21 days and 28 days of initial diagnosis, respectively (**Figure 2c**). Strikingly, in both cases the majority of these patients are not hospitalized; i.e. 158 of 231 patients (68.4%, in the 21+ days category) and 61 of 99 patients (61.6%, in the 28+ days category).

Of 104 COVID_pos_ patients with available SARS-CoV-2 IgG serology data, 14 patients remain either serology negative or indeterminate till date. Of the remnant 90 patients who are seroconverted till date, the upper bound of time to IgG-seropositivity from initial PCR diagnostic testing has a mean of 37.8 days (95% confidence interval: 34.3-41.3 days; **Figure 3a**). Here, we consider this analysis an “upper bound”, rather than a definitive time to seropositivity, as each patient may have experienced IgG seroconversion prior to the date when the serology test was actually administered. Based on the limited longitudinal real-world data available for IgG seropositivity, this upper bound is the best estimate we are able to obtain at this juncture.

**Figure 3.**
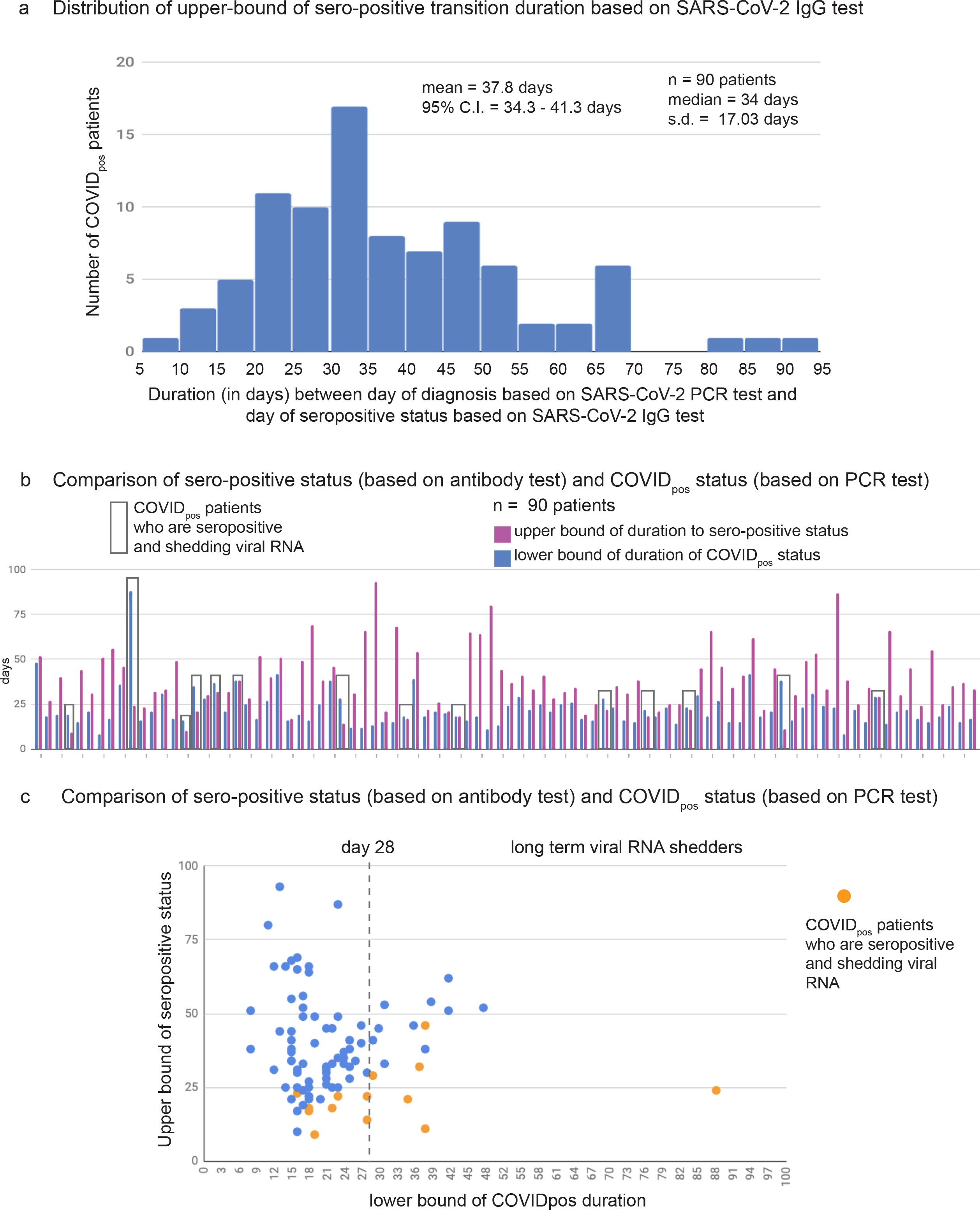
Distribution of upper-bound of the duration to sero-positive status based on SARS-CoV-2 IgG test and comparison to COVID_pos_ status based on SARS-CoV-2 PCR test. **(a)** Histogram of duration (in days) between the day of diagnosis based on SARS-CoV-2 PCR test and day of seropositive status based on SARS-CoV-2 IgG test. **(b)** Comparison of sero-positive status (based on antibody test) and COVID_pos_ status (based on PCR test). Cases that are both IgG-sero-positive and PCR positive are boxed. **(c)** Scatter plot of lower bound of viral RNA shedding (x-axis) versus the upper bound of IgG-seropositivity status (y-axis).

Next, we juxtaposed the SARS-CoV-2 PCR results against the IgG-seropositivity results for 90 COVID-19 patients with both sets of longitudinal data (**Figure 3b**). Surprisingly, 14 of these patients had viral RNA shedding between 0 to 64 days after their confirmed date of IgG-seropositivity (**Figure 3c**). The finding that the time to IgG seropositivity can be shorter than the lower bound of positive PCR tests in some patients, suggests that COVID-19 patients can continue to shed viral RNA for days or even weeks while generating IgG antibodies.

Long-term SARS-CoV-2-positive PCR tests are *not* necessarily indicative of long-term replication-competent virus.^12,13^ Consequently, we evaluated 488 SARS-CoV-2-positive PCR test’s Crossing point (Cp) values from nasopharyngeal swab samples of 208 patients. Among these, the mean Cp value was 28.6 with a standard deviation of 5 (**Figure 4a**). From 63 patients with viral RNA shedding beyond 21 days of initial diagnosis, the distribution of Cp values showed a mean of 31.87 and standard deviation of 1.14 (**Figure 4b**). In a subset of 17 patients who demonstrated longer-term viral RNA shedding beyond 28 days of initial diagnosis, the Cp values had a mean of 31.93 with a standard deviation of 1.22 (**Figure 4c**). The overall decreased mean Cp value across all compiled positive PCR tests relative to the long-term shedder’s PCR test Cp values, implies a smaller amount of viral RNA available for amplification in the swab samples from long-term shedders (post 21 and 28 days of initial diagnosis).

**Figure 4.**
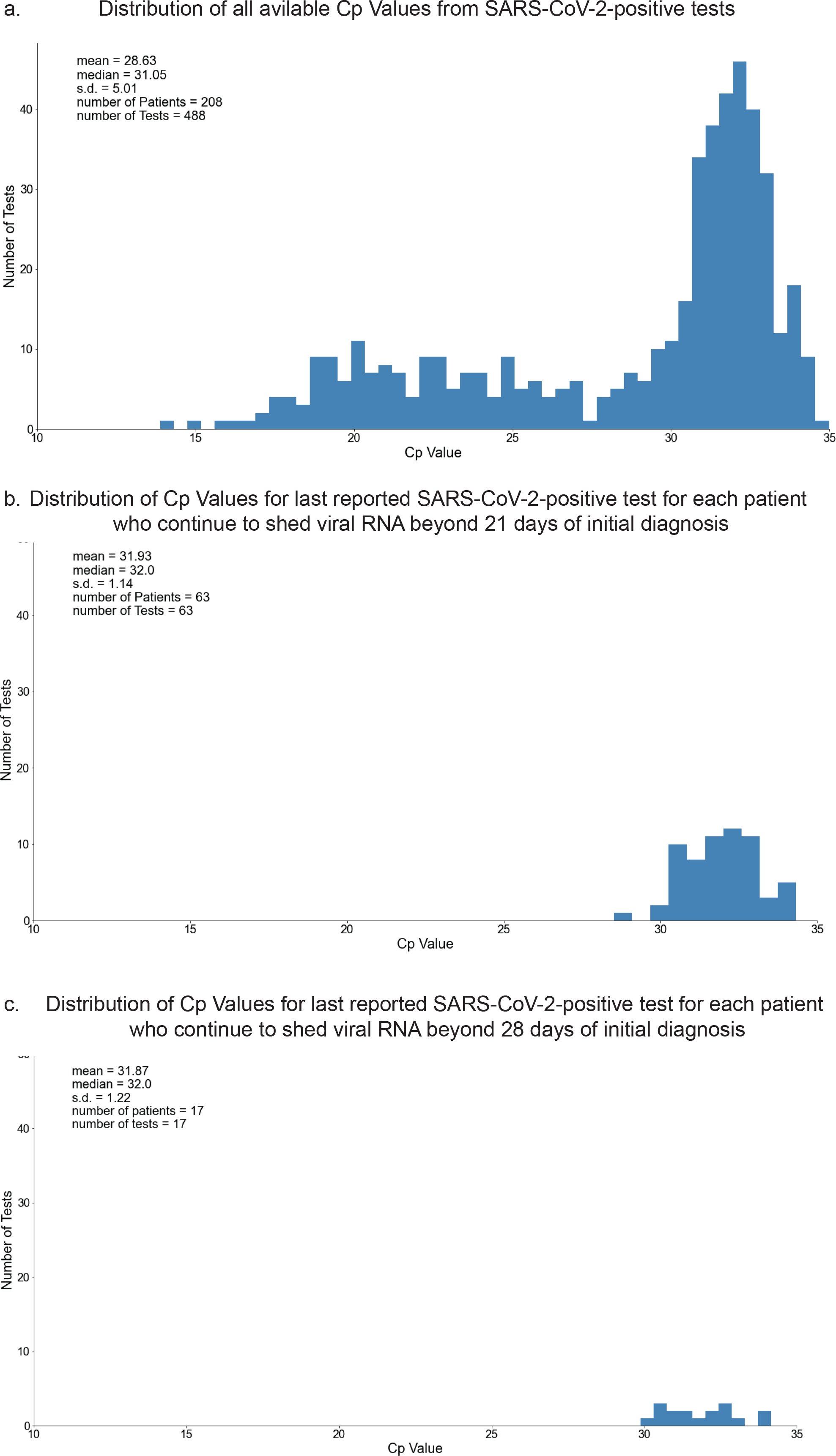
Distributions of RT-PCR Crossing point (Cp) values: **(a)** all reported SARS-CoV-2-positive tests; **(b)** the last reported SARS-CoV-2-positive test for each patient who continue to shed viral RNA beyond 21 days of initial diagnosis; and **(c)** the last reported SARS-CoV-2-positive test for each patient who continue to shed viral RNA beyond 28 days of initial diagnosis.

## Discussion

A recent study from China of 74 COVID_pos_ patient’s fecal samples and respiratory swabs observed SARS-CoV-2-positive swabs with a mean duration of 15.4 days and standard deviation of 6.7 days from the first symptom onset^15^. In this study, we have shown that COVID-19 patients that are long-term shedders are predominantly not hospitalized, thus underlining the importance of understanding the temporal dynamics of viral load, the duration of infectivity, and the likelihood of community transmission. Studies focusing on the temporal profiles of viral shedding suggest that the viral loads are highest at symptom onset, and subsequently decrease over the following 21 day^13,16^ and that live virus could no longer be cultured after day 8, leading to the hypothesis that SARS-CoV-2 infectiousness may decline from the time of symptom onset^12,13^. Whether such experimental results are generalizable to all COVID-19 patients that are long-term shedders is an important follow-up question.

Based on a recent study that characterized SARS-CoV-2 viral replication as a function of RT-PCR Cp values, attempting a characterization of 488 PCR tests from 208 SARS-CoV-2-positive patients in our study,^7^ would suggest percentage of positive cultures of mean 49.2% and standard deviation 23.7% (**Figure 4 - Supplementary Figure 1a**). Likewise, extrapolating the corresponding distribution of percentage of positive culture values in the 63 patients with viral RNA shedding beyond 21 days based on the last positive PCR tests would have mean of 33.3% and standard deviation of 7.6% (**Figure 4 - Supplementary Figure 1b**), while these figures beyond the 28 days threshold would have mean of 33.6% and standard deviation of 8.2% (**Figure 4 - Supplementary Figure 1c)**. Given it is impossible to effectively apply the relationship garnered between Cp values and positive cultures from different published RT-PCR assays towards this study, positive culture characterization for our RT-PCR assay would involve a follow-up investigation that incorporates longitudinal viral load assessments.

Although the SARS-CoV-2-positive RT-PCR tests by no means causally implicate replication-competent virus, the presence of viral RNA for several weeks from initial infection certainly warrants longitudinal monitoring of the viral load^5,7^. Nonetheless, the question still remains as to why some COVID_pos_ patients harbor virus or viral RNA for far longer than other COVID_pos_ patients. Our observations regarding non-hospitalized, IgG-seropositive, long-term viral RNA shedders highlights the need for follow-up studies to study the neutralizing potential for antibodies generated by long-term shedders and contextualize such analysis vis-a-vis direct assessment of viral loads and more quantitative assays such as ddPCR.^5,6^

The assessment of whether any of the IgG and IgM antibodies generated are able to neutralize the SARS-CoV-2 surface proteins (spike, envelope, membrane) or the nucleocapsid protein^14^ would add an immunological lens to interpret the seroconversion upper bounds noted in this study. Taken together, such additional research would help inform whether current CDC guidelines of 10 days self-quarantining for asymptomatic patients may be broadly satisfactory, including for patients that are noted to be long-term SARS-CoV-2 RNA shedders^8^.

Several factors could influence the persistent SARS-CoV-2 PCR positive status. Replicative fitness of a given virus is one of them. For instance, in HIV, not all viruses replicate equivalently, and differences may be attributable in part to polymorphisms in different genes^18^. For SARS-CoV-2, there are reports of different polymorphisms that have been speculated to impact disease severity or transmissibility, such as the D164G mutation in the spike (S) protein^19^. Another factor that may influence the persistence of PCR positivity could be the timing and robustness of the immune response. For example, given the role of the IFN response in viral shedding^20^, early IFN response is likely to be beneficial and reduce shedding, whereas late IFN response may be deleterious and delay clearance. Another potential factor to consider is the T cell response^21^. When T cells express high levels of different effector mechanisms (e.g. Perforin/granzyme B, IFN), they are thought to work better than if they produce only one effector pathway.

It may be noted that the clinical sensitivity of SARS-CoV-2 PCR tests has been debated to some extent^17^, and certainly there are anecdotal examples from our own clinic experience where critical ill COVID-19 patients can switch from a COVID_pos_ status to a COVID_neg_ status within a short period of time. To robustly enable scientific assessment of the sensitivity of the routine RT-PCR testing data analyzed here, we summarized the entire pattern of serial PCR outcomes across the 5,569 COVID-19 patients in this study. This analysis shows that the vast majority of the COVID-19 patients subjected to our RT-PCR assays do produce consistent results, as determined by multiple contiguous PCR tests resulting in the same outcome. There is a small minority of COVID-19 patients where aberrant switching of PCR results is indeed observed, with the underlying reasons undetermined at this juncture.

In order to understand whether long-term SARS-CoV-2 RNA shedders display any other distinctive features from the structured EHR, we defined a control cohort of COVID-19 patients with an *upper bound of infection duration* between 1 to 13 days (“short-term shedders”). We compared the long-term shedders with this control cohort by analyzing the counts of over 15,000 features constituting the fields of structured EHR databases, including diagnosis, ICD codes, medication history, immunization records, procedures, and demographics (*see Methods*). We do not find any significant distinguishing clinical features for long-term shedders compared to short-term shedders across multiple scenarios: (1) all patients, features from last 90 days, (2) all patients, features from after diagnosis, (3) all patients, features from 21 days after diagnosis, (4) all patients, features from 28 days after diagnosis, (5) non-hospitalized patients, features from last 90 days, (6) non-hospitalized patients, features from 21 days after diagnosis, (7) non-hospitalized patients, features from 28 days after diagnosis, and (8) non-hospitalized patients, features from diagnosis date.

While this preliminary observation from structured EHR counts has to be monitored as more COVID-19 patients’ longitudinal lab results are available, at this juncture, it appears that the majority of long-term shedders are non-hospitalized patients with likely mild or moderate symptoms that do not prompt their detailed clinical follow-up. Given that some of these long-term viral RNA shedders are already seropositive when they continue shedding, suggests that we cannot yet rely on IgG or any other antibody response alone to estimate immunity or odds of re-infection potential with SARS-CoV-2 without investigating the salience of long-term RNA shedding. The fact that replication-competent virus in some of these long-term RNA shedders cannot be ruled out based on the data available from PCR Ct values, underlines the need for prolonged monitoring of viral loads and immune responses in such COVID-19 patients.

Our findings raise important additional follow-up questions from the standpoint of analyzing actual complete lab testing results longitudinally for large COVID-19 patient cohorts. Recent studies have identified coagulation associated issues in COVID-19 patients^22,23^. Building on findings reported in this study, it may be interesting to examine the rate of change of coagulation signals (e.g. by longitudinal lab testing of platelet count, fibrinogen levels, d-dimer values) and the levels of immune cells (e.g. neutrophils, monocytes, basophils, lymphocytes) in COVID-19 patients that are long-term SARS-CoV-2 RNA shedders versus patients who are able to more rapidly eliminate the viral RNA. Follow up studies need to examine how the duration of SARS-CoV-2 PCR positive status correlates to the rate of IgG seroconversion and the presence of effective humoral immunity as measured by neutralizing antibodies.

## Methods

### SARS-CoV-2 diagnostic tests conducted by Mayo Clinic hospitals and health system

Patients seen at Mayo Clinic in Rochester MN were tested by either a laboratory-developed test or the Roche cobas SARS-CoV-2 assay^25,26^. The Roche cobas test was employed by the Mayo Clinic’s Florida hospitals, and the Abbott diagnostic test was used by the Mayo Clinic’s Arizona hospitals^27^. These SARS-CoV-2 PCR tests amplify different segments of the viral genome but are considered largely equivalent from the perspective of their analytical performance. The Logical Observation Identifiers Names and Codes (LOINC) code of the SARS-CoV-2 IgG test analyzed is 94563-4 (https://loinc.org/94563-4/)^28^.

### Statistical analysis of longitudinal SARS-CoV-2 RT-PCR results

The features considered in the analysis to differentiate the COVID-19 patients that are persistently PCR positive include all structured entities from the EHR, including but not limited to demographics, diagnosis, International Statistical Classification of Diseases and Related Health Problems (ICD) codes, medication history, immunization record, procedures, vitals and lab tests. Any feature which is enriched significantly towards either shorter durations (less than 14 days between first positive to second negative test, as depicted in **Figure 2b**) or longer durations (greater than or equal to 28 days between first positive to recent/final positive test, as depicted in **Figure 2c**) was noted down. During the observation period (n = 201; 99 persistently PCR positive patients; 102 control patients), there were 289 EHR-derived features that were considered, including potentially prior to each patient’s COVID-19 diagnosis. The 2-proportion z-test p-value (after Benjamini-Hochberg [BH] adjustment for multiple hypothesis correction) was used to assess the differences of each feature between the persistently PCR positive patients and the control cohort, defined as those COVID-19 patients with an *upper bound of infection duration* between 1 to 13 days. The procedure was as follows:

1. Filter by features which are present in overall at least 10% of the patients we’re looking at.
2. It’s possible that there is a bias of more overall features towards the long-term or control cohort. We are not interested in this bias. To account for this, for each feature, we compute the “baseline” proportion difference, i.e. the weighted mean proportion of persistently positive patients that are positive for that feature minus the weighted mean proportion of control cohort which is positive for that feature. Call this baseline difference *O* (we have one such *O* for each feature).
3. Perform a 2-proportion *z*-test for whether the difference between feature-positive rate in the long-term cohort and feature-positive rate in the control cohort is significantly different from the baseline *O*.
4. Adjust these *p*-values for multiple hypotheses using the Benjamini-Hochberg procedure (with False Discovery Rate [FDR] controlled at 0.1 level).

We repeated the above procedure for slightly different underlying data as well; in particular, we re-ran on the following variants:

i. We filtered to look only at patients who were not hospitalized (as those would be of most concern).
ii. Each binary feature (phenotype, lab test, etc) occurred at a particular day in the patient’s record. We filtered by only those features which occur 0, 21 or 28 days following diagnosis.
iii. Variations (i) and (ii) together

### Prediction of Positive Culture Probability by RT-PCR Crossing point (Cp) Value

Estimated probability of positive culture (‘y’) by RT-PCR cycle threshold Crossing point (Cp) values (‘x’) was calculated using the following formula.

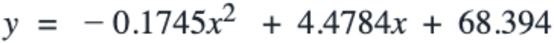

This formula was derived by fitting a quadratic equation to RT-PCR Cp Values and positive culture rates, as reported by La Scola, B. et al^7^.

## Data Availability

Please contact the corresponding author regarding data availability. Please review HIPAA regulations and Mayo Clinic IRB guidelines before making any requests.

## Acknowledgments

The authors thank Mathai Mammen, Murali Aravamudan, Patrick Lenehan, Will Gibson, Jacob Martin, Travis Hughes, and Tyler Wagner for their helpful feedback on this research.

## Figure Legends

**Figure 4 –Supplementary Figure 1.**
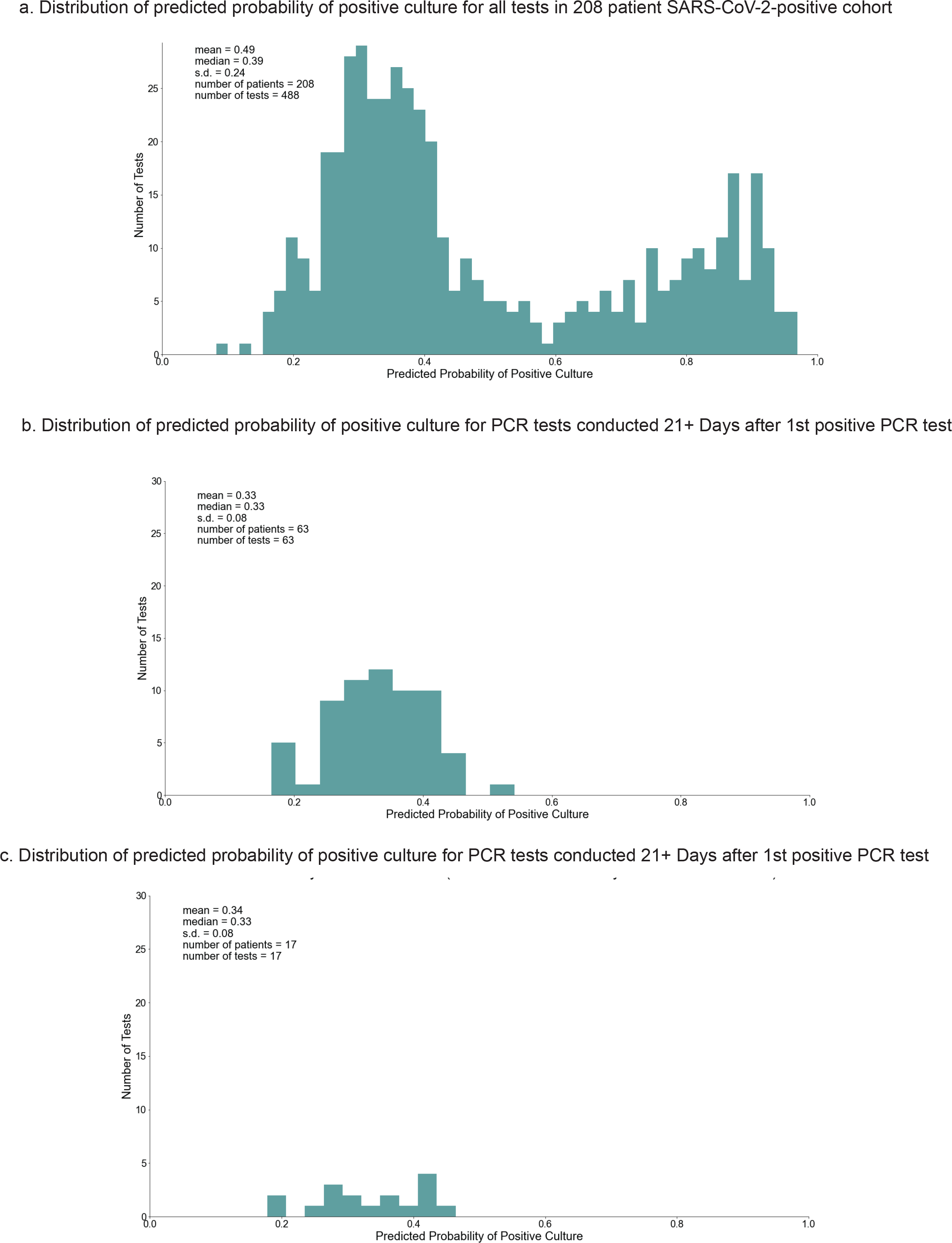
Extrapolating the distribution of percentage of positive culture values from PCR Ct values, for: **(a)** all tests from 208 COVID-19 patients, **(b)** tests conducted after 21 days of initial diagnosis, **(c)** tests conducted after 28 days of initial diagnosis.

